# Agent Orange exposure and prostate cancer risk in the Million Veteran Program

**DOI:** 10.1101/2023.06.14.23291413

**Authors:** Asona J. Lui, Meghana S. Pagadala, Allison Y. Zhong, Julie Lynch, Roshan Karunamuni, Kyung Min Lee, Anna Plym, Brent S. Rose, Hannah Carter, Adam S. Kibel, Scott L. DuVall, J. Michael Gaziano, Matthew S. Panizzon, Richard L. Hauger, Tyler M. Seibert

## Abstract

**Purpose:** Exposure to Agent Orange, a known carcinogen, might increase risk of prostate cancer (PCa). We sought to investigate the association of Agent Orange exposure and PCa risk when accounting for race/ethnicity, family history, and genetic risk in a diverse population of US Vietnam War veterans.

**Methods & Materials:** This study utilized the Million Veteran Program (MVP), a national, population-based cohort study of United States military veterans conducted 2011-2021 with 590,750 male participants available for analysis. Agent Orange exposure was obtained using records from the Department of Veterans Affairs (VA) using the US government definition of Agent Orange exposure: active service in Vietnam while Agent Orange was in use. Only veterans who were on active duty (anywhere in the world) during the Vietnam War were included in this analysis (211,180 participants). Genetic risk was assessed via a previously validated polygenic hazard score calculated from genotype data. Age at diagnosis of any PCa, diagnosis of metastatic PCa, and death from PCa were assessed via Cox proportional hazards models.

**Results:** Exposure to Agent Orange was associated with increased PCa diagnosis (HR 1.04, 95% CI 1.01–1.06, p=0.003), primarily among Non-Hispanic White men (HR 1.09, 95% CI 1.06– 1.12, p<0.001). When accounting for race/ethnicity and family history, Agent Orange exposure remained an independent risk factor for PCa diagnosis (HR 1.06, 95% CI 1.04–1.09, p<0.05). Univariable associations of Agent Orange exposure with PCa metastasis (HR 1.08, 95% CI 0.99–1.17) and PCa death (HR 1.02, 95% CI 0.84–1.22) did not reach significance on multivariable analysis. Similar results were found when accounting for polygenic hazard score.

**Conclusions:** Among US Vietnam War veterans, Agent Orange exposure is an independent risk factor for PCa diagnosis, though associations with PCa metastasis or death are unclear when accounting for race/ethnicity, family history, and/or polygenic risk.

## Introduction

Agent Orange, a mixture of herbicides used predominately in the Vietnam war from August 1965 to February 1971 to clear dense vegetation and destroy food crops (Operation Ranch Hand), was contaminated with a dioxin compound known as 2,3,7,8-tetrachlorobenzo-p-dioxin (TCDD).^1^ TCDD is absorbed quickly, eliminated slowly, and has a half-life of 7.2 years in the human body.^2-4^ It also causes direct DNA damage and alterations in growth factor signaling which cause aberrant cell growth and tumors in exposed persons.^1,5^ TCDD has been classified as a carcinogen since the 1990s and has been definitively linked to soft tissue and hematological malignancies in exposed Vietnam war veterans, which is defined by the Agent Orange act of 1991 to be all Veterans who served anywhere in Vietnam between January 9, 1962 to May 7,1975.^6-8^ However, it was not until the 2000s that a potential association between Agent Orange exposure in the Vietnam War and genitourinary cancers was acknowledged.^1^ Since then, evidence linking Agent Orange exposure to increased PCa risk or associated mortality among Vietnam War veterans has been limited to small case series.^9-16^ These small studies have found a slightly lower age at diagnosis, higher incidence of Stage IV disease and lower rates of biochemical control, though differences in overall survival or other clinical outcomes have not been found.^9,16,17^

Meanwhile, PCa risk in the general population is associated most strongly with age, race/ethnicity (especially Black race), family history, and specific genetic elements.^17-23^ We sought to investigate the association between Agent Orange exposure and PCa risk in a large, diverse population when accounting for race/ethnicity, family history, and genetic risk. We used data from the Department of Veteran Affairs (VA) Million Veteran Program (MVP), a population-based cohort with genotyping, long-term follow-up, and linked clinical records for over 870,000 participating US veterans. The MVP is one of the largest and most diverse electronic health record-linked biobanks in the world, with a unique structure that allows for detailed investigation into the interactions between inherited risk and Agent Orange exposure in 211,180 US veterans.^24^ We tested Agent Orange exposure for associations with age at PCa diagnosis, age at diagnosis of PCa metastasis, and lifetime PCa-specific mortality.

## Methods

### Participants

We obtained data from MVP for individuals recruited from 63 Veterans Affairs Medical Centers across the United States (US) beginning in 2011. All veterans were eligible for participation in MVP. Study participation included consenting to access the participant’s electronic health records for research purposes. The MVP received ethical and study protocol approval from the VA Central Institutional Review Board in accordance with the principles outlined in the Declaration of Helsinki. Only men were included in the present study of PCa risk. Further, we limited the present study to those who were on active duty during the Vietnam War era (1955-1975). Participant characteristics are shown in **Table 1**.

**Table 1:** Participant characteristics for self-reported race/ethnicity groups during the Vietnam War. Numbers indicate the number of participants available for analysis.

|  | All | Self-reported Race/Ethnicity |  |  |  |  |  |  |  |
| --- | --- | --- | --- | --- | --- | --- | --- | --- | --- |
|  |  | Non-Hispanic White | Black or African American | Hispanic White | Asian | Native American | Pacific Islander | Other | Unknown |
| Active duty during Vietnam War | 211,180 | 172,170 | 23,923 | 6,370 | 1,450 | 2,279 | 376 | 4,100 | 512 |
| Fatal prostate cancer | 518 | 387 | 100 | 14 | <10 | <10 | <10 | 13 | <10 |
| Metastatic prostate cancer | 2,445 | 1,766 | 511 | 75 | 17 | 18 | <10 | 47 | <10 |
| Any prostate cancer | 29,667 | 22,303 | 5,559 | 796 | 170 | 266 | 41 | 479 | 53 |

### Potential Agent Orange Exposure

Potential exposure to Agent Orange was determined by the VA Compensation & Pension Committee, as recorded in the MVP data core. Per the legal US government definition, Veterans who served physically (on land or inland waterways) in Vietnam during periods of Agent Orange use by the US military were considered exposed to Agent Orange (January 9,1962-May 7,1975). Information about the intensity (amount and duration) of Agent Orange exposure for each individual is not known.

### Clinical Data Extraction

Comprehensive description of clinical endpoints used for PCa Cox proportional hazards analysis were described previously.^17^ Briefly, PCa diagnosis, age at diagnosis, PSA tests, and date of last follow-up were retrieved from the VA Corporate Data Warehouse based on ICD codes and VA Central Cancer Registry data. Age at diagnosis of metastatic PCa indicated the age of the participant when diagnosed with either nodal or distant metastases as determined through a validated natural language processing tool.^17,25^ Fatal PCa information was determined from National Death Index. Participants with ICD10 code “C61” as underlying cause of death were considered to have died from PCa. Family history was recorded as either the presence or absence of (one or more) first-degree relatives with PCa. Among the participants eligible for analysis, over 99% had received at least one PSA test in the VA system, though the age at testing and frequency of testing were variable and clinical indications (screening vs. diagnostic workup) are not known.

### Genetic Risk: Polygenic Hazard Score (PHS290)

Blood sampling and DNA extraction were conducted by MVP as described previously.^1,14^ The MVP 1.0 genotyping array contains a total of 723,305 variants, enriched for low frequency variants in African and Hispanic populations and variants associated with diseases common to the VA population.^15^ Details on quality control and imputation have been described previously.^14^

To assess genetic risk, we calculated a previously developed and validated polygenic hazard score using 290 common genetic variants (PHS290) that reliably stratifies men for age-dependent genetic risk of PCa and is associated with PCa, metastatic PCa, and PCa death.^17,21^ Details of PHS290 calculation in MVP are described elsewhere.^17,21^ PHS290 performs well in diverse datasets and is independently associated with PCa risk.^17,21,26^

### Cox Proportional Hazards Analysis

We used Cox proportional hazards models to evaluate the association of Agent Orange exposure with three clinical endpoints: age at diagnosis of PCa, age at diagnosis of metastatic PCa, and age at death from PCa. We also analyzed self-reported racial/ethnic subgroups. As very few participants identified as being of both Black race and Hispanic ethnicity, we included these in a single category for Black or African-American race. Where individuals did not meet the endpoint of interest, we censored at age at last follow-up.

To assess for independent association of Agent Orange exposure with PCa endpoints, we used multivariable Cox proportional hazards models. We tested Agent Orange status associations in models with race/ethnicity and family history, without and with PHS290. For race/ethnicity hazard ratios, we used Non-Hispanic White as reference. For PHS290, we illustrated the effect size via the hazard ratio for the highest 20% vs. lowest 20% of genetic risk (HR80/20) and between other strata of PHS290. These percentiles refer to previously defined thresholds of PHS290 derived from European men unaffected by PCa and <70 years old. Further details and assessment of alternate strategies in the MVP population is described elsewhere.^17,21^ We assessed statistical significance with two-tailed alpha at 0.01.

### PSA testing

Screening has been shown in a large, randomized trial to increase PCa incidence and reduce cause-specific mortality^27^, raising the possibility that PSA testing may confound any impact of Agent Orange exposure. The reason for PSA tests in MVP participants is unknown, but we were able to count the number of PSA tests each participant underwent. We tested for associations between potential Agent Orange exposure and number of pre-diagnostic PSA tests (i.e., ≥2 years prior to a PCa diagnosis) for a given participant by performing univariable and multivariable linear regressions. Multivariable linear regressions used race/ethnicity, family history, and PHS290 as predictive variables in addition to Agent Orange exposure. Prior analysis of MVP data showed that increased pre-diagnostic PSA testing was associated with decreased risk of PCa, a counterintuitive result explained by the fact that men never diagnosed with PCa were eligible for more pre-diagnostic PSA tests. No such confound exists for those men never diagnosed with PCa, however, and all PSA tests in this group are pre-diagnostic. Therefore, if Agent Orange exposure leads to more frequent PSA testing, we would expect to observe a positive association of Agent Orange and number of PSA tests among these participants.

## Results

Median age at MVP enrollment was 67 years (interquartile range 64-70). Median age at last follow-up was 72 (69-74).

On univariable analysis, Agent Orange exposure was associated with increased PCa diagnosis (HR 1.04, 95% CI 1.01–1.06, p=0.003) (**Table 2**), primarily driven by the association in the large Non-Hispanic White subgroup (HR 1.09, 95% CI 1.06–1.12, p<0.001). Agent Orange exposure was was not associated with increased risk of metastatic PCa among all participants, but there was a significant association in only the Non-Hispanic White subgroup (HR 1.17, 95% CI 1.06– 1.3, p=0.002). In Black and Hispanic White participants, there was no evidence of association between Agent Orange exposure and PCa risk (HR 1.01, 95%CI 0.95–1.07, p=0.858 and HR 0.99, 95% CI 0.85–1.15, p=0.937). Cause-specific cumulative incidence curves for PCa were qualitatively fairly similar regardless of Agent Orange exposure status (**Figure 1**).

**Table 2:** Univariable association of Agent Orange exposure with any, metastatic and fatal PCa. Hazard ratios (HR) from Cox Proportional Hazards model of Agent Orange exposure with age at prostate cancer, metastatic prostate cancer (nodal or distant) and death from prostate cancer. *P*-values reported are from univariable models using Agent Orange exposure (yes or no) as the sole predictor variable. Significant associations are indicated by *. NA, indicates subgroups for which there were insufficient participants to estimate results. n, indicates number at risk.

| Race/Ethnicity | Any Prostate Cancer |  | Metastatic Prostate Cancer |  | Fatal Prostate Cancer |  |
| --- | --- | --- | --- | --- | --- | --- |
|  | HR | P | HR | P | HR | P |
| All<br>(n=211,180) | 1.04 [1.01-1.06] | 0.003* | 1.05 [0.97-1.15] | 0.232 | 1.01 [0.82-1.22] | 0.945 |
| Non-Hispanic White<br>(n=172,170) | 1.09 [1.06-1.12] | <0.001* | 1.17 [1.06-1.3] | 0.002* | 1.1 [0.88-1.34] | 0.401 |
| Black or African American<br>(n=23,923) | 1.01 [0.95-1.07] | 0.858 | 0.86 [0.69-1.04] | 0.142 | 0.9 [0.57-1.38] | 0.654 |
| Hispanic White<br>(n=6,370) | 0.99 [0.85-1.15] | 0.937 | 0.84 [0.49-1.32] | 0.488 | 0.6 [0.0-1.99] | 0.438 |
| Asian<br>(n=1,450) | 0.68 [0.47-0.97] | 0.054 | 0.92 [0.17-2.42] | 0.891 | NA | NA |
| Native American<br>(n=2,279) | 1.31 [1.02-1.65] | 0.038 | 0.89 [0.2-2.37] | 0.831 | NA | NA |
| Pacific Islander<br>(n=376) | 1.25 [0.66-2.35] | 0.494 | NA | NA | NA | NA |
| Other<br>(n=4,100) | 0.91 [0.75-1.1] | 0.330 | 0.88 [0.43-1.63] | 0.689 | 0.4 [0.0-1.35] | 0.237 |
| Unknown<br>(n=512) | 0.85 [0.44-1.49] | 0.586 | NA | NA | NA | NA |

**Figure 1.**
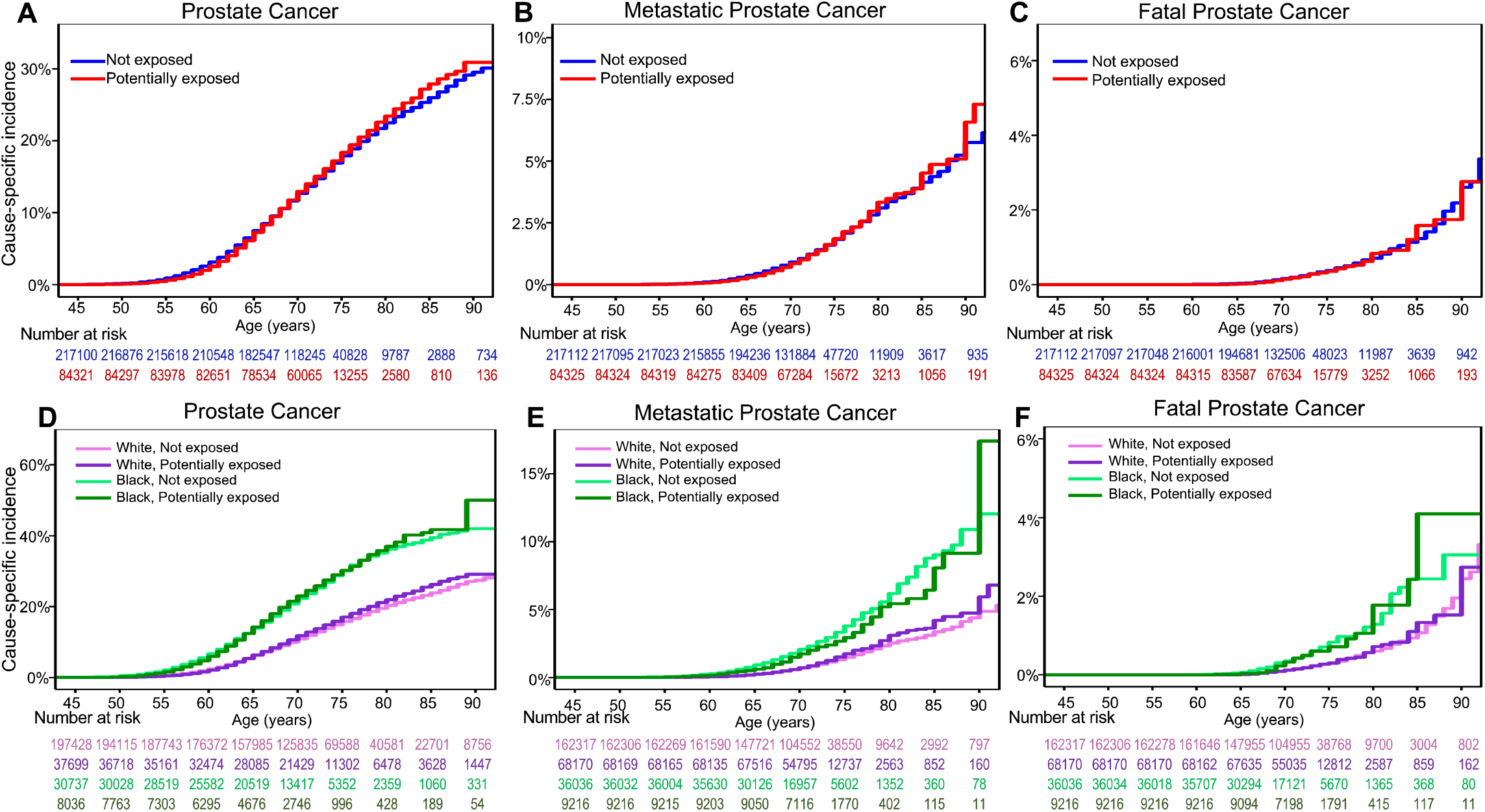
Million Veteran Program (MVP) Cause-specific Cumulative Incidence based on Agent Orange exposure. Cause-specific cumulative incidence among MVP participants on active duty during the Vietnam War, stratified by Agent Orange exposure status (top row) and stratified by self-reported race (bottom row) for **(A, D)** all prostate cancer, **(B, E)** metastatic prostate cancer, and **(C, F)** fatal cancer. “White” indicates Non-Hispanic White participants, and “Black” indicates Black and Hispanic Black participants.

When accounting for race/ethnicity and family history, Agent Orange exposure remained an independent risk factor for PCa diagnosis (HR 1.06, 95% CI 1.04–1.09, p<0.05) but not for metastatic PCa or PCa death. (**Table 3**). Accounting for genetic risk (PHS290) as well as race/ethnicity and family history had no impact on the association between Agent Orange exposure status and PCa diagnosis (HR 1.06, 95% CI 1.04–1.09, p<0.05) (**Table 4**). Agent Orange exposure status did not differentially modulate absolute PCa risk among men with high genetic risk (PHS290 >80^th^ percentile, as defined previously^21^) or across any PHS290 values (**Supplemental Figure 1 and Supplemental Table 1**).

**Table 3:** Multivariable models combining self-reported Race/Ethnicity, Family History and Agent Orange Exposure for three PCa clinical endpoints. Cox proportional hazards results for association with age at death from PCa, at diagnosis of metastatic PCa and age at diagnosis with PCa. *P*-values reported are from multivariable models using self-reported race/ethnicity, family history and Agent Orange exposure (yes or no). Hazard ratios for race/ethnicity were estimated using Non-Hispanic White as the reference. Hazard ratios for family history were for one or more first-degree relatives diagnosed with prostate cancer. This multivariable analysis was limited to the 211,180 participants who were on active duty during the Vietnam War. Numbers in brackets are 95% confidence intervals. Significant predictors in the multivariable model are indicated by *(*p*<0.05), **(p<0.01) and *** (*p*<0.001).

| Clinical Endpoint | Self-Reported Race/Ethnicity |  |  |  |  |  |  | Family History | Agent Orange Exposure |
| --- | --- | --- | --- | --- | --- | --- | --- | --- | --- |
|  | Black or African American | Hispanic White | Asian | Native American | Pacific Islander | Unknown | Other |  |  |
| Fatal Prostate Cancer | 2.37 [1.87-2.92]*** | 1.09 [0.58-1.65] | 0.27 [0.0-0.84] | 0.79 [0.0-1.82] | 0.0 [0.0-0.0] | 0.0 [0.0-0.0] | 2.06 [1.01-3.28] | 1.92 [1.47-2.39]*** | 1.02 [0.84-1.22] |
| Metastatic Prostate Cancer | 2.51 [2.27-2.75]*** | 1.23 [0.97-1.55] | 1.05 [0.58-1.6] | 0.94 [0.51-1.38] | 0.57 [0.0-1.52] | 1.86 [0.79-3.22] | 1.46 [1.07-1.92] | 1.5 [1.33-1.7]*** | 1.08 [0.99-1.17] |
| Prostate Cancer | 2.2 [2.14-2.27]*** | 1.02 [0.95-1.1] | 0.88 [0.75-1.02] | 1.02 [0.91-1.15] | 0.91 [0.65-1.21] | 0.84 [0.62-1.08] | 1.06 [0.96-1.16] | 1.86 [1.8-1.92]*** | 1.06 [1.04-1.09]* |

**Table 4:**
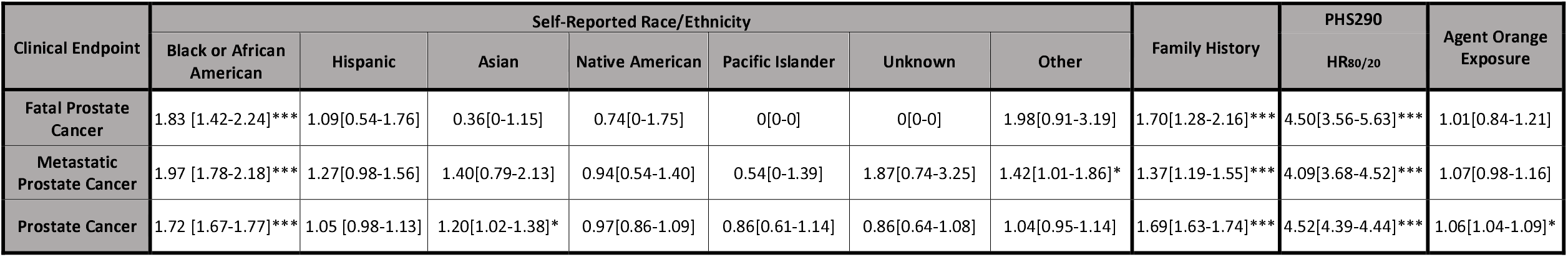
Multivariable models combining self-reported Race/Ethnicity, Family History, genetic risk and Agent Orange Exposure for three PCa clinical endpoints. Cox proportional hazards results for association with age at death from PCa, at diagnosis of metastatic PCa and age at diagnosis with PCa. *P*-values reported are from multivariable models using self-reported race/ethnicity, family history, genetic risk (PHS290) and Agent Orange exposure (yes or no). For PHS290, effect size was illustrated via the hazard ratio (HR_80/20_) for the highest 20% vs. lowest 20% of genetic risk. Hazard ratios for race/ethnicity were estimated using Non-Hispanic White as the reference. Hazard ratios for family history were for one or more first-degree relatives diagnosed with prostate cancer. This multivariable analysis was limited to the 211,180 participants who were on active duty during the Vietnam War. Numbers in brackets are 95% confidence intervals. Significant predictors in the multivariable model are indicated by *(*p*<0.05), **(p<0.01) and *** (*p*<0.001).

To assess whether differences in PSA testing account for the association between Agent Orange exposure and PCa risk, we conducted univariable and multivariable linear regressions on Vietnam era MVP participants who never developed PCa. On average, men in this population received approximately 8 PSA tests in the VA system (Table 5). Agent Orange exposure resulted in a statistically significant but small reduction in screening intensity (7.4 PSA tests) on univariable analysis. Several variables were independently associated with the number of PSA tests reported per MVP participant, including participant race and family history, concordant with guidelines that support stronger consideration of screening for men at higher risk^28^ (**Table 5**). Agent Orange exposure was also an independent predictor of a small reduction in PSA testing (7.4 PSA tests) on multivariable linear regression. Therefore, the increase in PCa diagnosis associated with Agent Orange exposure is likely not due to increased screening intensity.

**Table 5:** Univariable and multivariable linear regressions assessing association between Agent Orange exposure, self-reported Race/Ethnicity, genetic risk and Family History (FH), with screening intensity (total number of PSA screening tests) among Vietnam War veterans without PCa. Results for association between each factor and number of PSA tests recorded in the VA medical record for a given participant. *P*-values reported are from linear models using Agent Orange exposure code, self-reported race/ethnicity, family history, family history and genetic risk (PHS290). Coefficients for race/ethnicity were estimated using Non-Hispanic White as reference. Family history was defined as one or more first-degree relatives diagnosed with prostate cancer. For PHS290, effect size was illustrated via the hazard ratio (HR_80/20_) for the highest 20% vs. lowest 20% of genetic risk. Numbers in brackets are 95% confidence intervals. Significant predictors in each model are indicated by *(*p*<0.01), **(p<0.001). On univariable analysis, Agent Orange exposure was associated with an average of 0.56 fewer PSA tests in this population than performed in men not exposed to Agent Orange.

| Univariable |  | Multivariable |  |  |  |  |  |  |  |  |  |  |
| --- | --- | --- | --- | --- | --- | --- | --- | --- | --- | --- | --- | --- |
| Intercept | Agent Orange Exposure | Intercept | Self-Reported Race/Ethnicity |  |  |  |  |  |  | Family History | PHS290<br>HR<sub>80/20</sub> | Agent Orange Exposure |
|  |  |  | Black | Hispanic White | Asian | Native American | Pacific Islander | Other | Unknown |  |  |  |
| 7.93<br>[7.91-7.95]** | -0.56<br>-[0.61-0.52]** | 8.71<br>[8.13-9.29]** | 1.24<br>[1.17-1.32]** | 0.19<br>[0.06-0.32]* | -1.26<br>-[1.53-0.99]** | 0.63<br>[0.42-0.85]** | -0.47<br>[-1.0-0.05] | 0.28<br>[0.11-0.44]* | -1.79<br>-[2.23-1.34]** | 0.12<br>[0.03-0.20]* | -0.11<br>-[0.18-0.05] | -0.62<br>[0.01-0.21]** |

## Discussion

In a large, diverse, population-based cohort of US veterans, Agent Orange exposure was associated with a modest increase in PCa diagnosis in the full cohort. Agent Orange was not significantly associated with metastatic PCa diagnosis for the full cohort, though a significant association was observed in the self-reported Non-Hispanic White subgroup. We also present the first multivariable analysis in a population-based cohort to assess whether Agent Orange exposure was an *independent* risk factor for PCa outcomes when accounting for family history, ancestry, and/or genetic risk. We found that Agent Orange exposure was associated only with PCa diagnosis and not with metastasis or PCa death. Our finding that Agent Orange exposure is associated more with PCa diagnosis than with aggressive cancer outcomes may be reassuring when counseling men in this population.

Details confirming actual Agent Orange exposure including duration or intensity are not available in MVP and are difficult to ascertain for patients in clinical practice, as well. Some veterans who physically served in Vietnam while Agent Orange was in use may have had heavy and/or frequent exposure, whereas others may have escaped with little to no exposure. It is possible that intense Agent Orange exposure is associated with aggressive PCa, though adequate data may never be available to answer this question. The definition of Agent Orange exposure using in this study (service in Vietnam during the years when Agent Orange was in widespread use) is also the US government’s official definition and is used by the VA Compensation & Pension Committee to address the needs of potentially exposed individuals. Using this definition estimates associations of the *average* exposure by those in Vietnam during use of Agent Orange and was sufficient to find significant associations with PCa, metastatic PCa, and fatal PCa on univariable analysis. Multivariable association analyses here may be underpowered to detect possible independent associations of Agent Orange with metastatic or fatal PCa. On the other hand, we can conclude that average Agent Orange exposure among Vietnam veterans has a much smaller effect size than do family history, Black race, or high polygenic risk. On multivariable analysis, potential Agent Orange exposure yielded estimated hazard ratios <1.10 for all PCa endpoints, whereas hazard ratios for metastatic PCa were 1.37 for family history, 1.97 for Black race, and 4.09 for polygenic risk (PHS290).

This study was conducted using the MVP dataset and so the results and conclusions may not be generalizable beyond the VA population. As sequencing for rare pathogenic mutations was not performed, it was also not possible to assess the impact of Agent Orange exposure on risk arising from, for example, germline *BRCA2* mutations. Potential differences in PCa screening intensity between exposure groups were not completely accounted for, though there was no evidence of increased PSA testing among those exposed to Agent Orange in this study.

## Conclusion

Among men who were on active duty during the Vietnam War, Agent Orange exposure (i.e., service in Vietnam while Agent Orange was in use) is identified as an independent risk factor for PCa diagnosis with small effect size. When accounting for family history, race/ethnicity, and polygenic risk, potential Agent Orange exposure was not independently associated with PCa metastasis or PCa death.

## Data Availability

It is not possible for the authors to directly share the individual-level data that were obtained from the MVP due to constraints stipulated in the informed consent. Anyone wishing to gain access to this data should inquire directly to MVP at. The data generated from our analyses are included in the manuscript main text, tables, and figures.

## Acknowledgements

This research used data from the Million Veteran Program, Office of Research and Development, Veterans Health Administration. This research was supported by the Million Veteran Program MVP022 award # I01 CX001727 (PI: Richard L. Hauger MD). This publication does not represent the views of the Department of Veterans Affairs or the United States Government.

## Table and Figure Legends

**Supplemental Figure 1.**
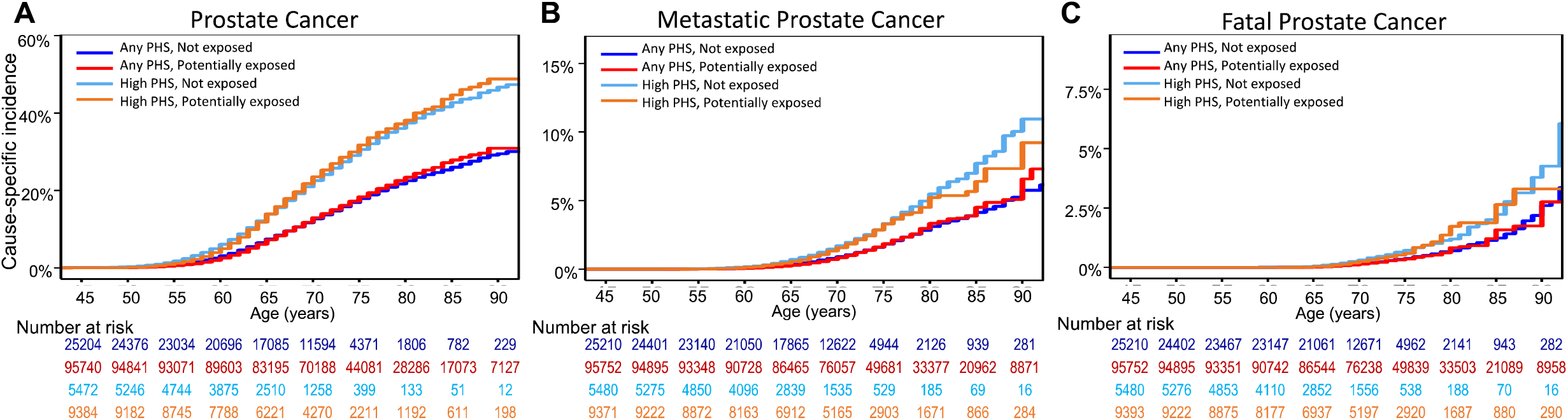
Million Veteran Program (MVP) Cause-specific Cumulative Incidence based on Agent Orange exposure for participants with high PCa risk. Cause-specific cumulative incidence among MVP participants on active duty during the Vietnam War, stratified by Agent Orange exposure and PHS290 for **(A)** all prostate cancer, **(B)** metastatic prostate cancer, and **(C)** fatal cancer. ‘High PHS290’ indicates participants with PHS in the top 20% of genetic risk, as assessed by PHS290, a polygenic hazard score.

**Supplemental Table 1:** Multivariable model with interaction term between Agent Orange exposure status and PHS290 for three PCa clinical endpoints. Multivariable linear regression results for an interaction between Agent Orange exposure code and PHS290 for age at death from PCa, at diagnosis of metastatic PCa and age at diagnosis with PCa. Coefficient for each interaction term ids displayed. *P*-values reported are from multivariable models using genetic risk (PHS290) and Agent Orange exposure (yes or no). Multivariable analysis was limited to the 211,180 participants who were on active duty during the Vietnam War.

| Clinical Endpoint | Coefficient | p-value |
| --- | --- | --- |
| Fatal Prostate Cancer | 0.0354 | 0.2927 |
| Metastatic Prostate Cancer | -0.1074 | 0.3519 |
| Prostate Cancer | -0.1031 | 0.6843 |

